# Association between live childhood vaccines and COVID-19 outcomes: a national-level analysis

**DOI:** 10.1101/2020.10.17.20214510

**Authors:** Chikara Ogimi, Pingping Qu, Michael Boeckh, Rachel A. Bender Ignacio, Sahar Z. Zangeneh

## Abstract

We investigated whether countries with higher coverage of childhood live vaccines [BCG or measles-containing-vaccine (MCV)] have reduced risk of COVID-19 related mortality, accounting for known systems differences between countries. In this ecological study of 140 countries using publicly available national-level data, higher vaccine coverage, representing estimated proportion of people vaccinated during the last 15 years, was associated with lower COVID-19 deaths. The associations attenuated for both vaccine variables, and MCV coverage became no longer significant once adjusted for a validated summary score accounting for life expectancy and healthcare quality indicators, the Healthcare access and quality index (HAQI). The magnitude of association between BCG coverage and COVID-19 death rate varied according to HAQI, and MCV coverage had little effect on the association between BCG and COVID-19 deaths. While there are associations between live vaccine coverage and COVID-19 outcomes, the vaccine coverage variables themselves were strongly correlated with COVID-19 testing rate, HAQI, and life expectancy. This suggests that the population-level associations may be further confounded by differences in structural health systems and policies. Cluster randomized studies of booster vaccines would be ideal to evaluate the efficacy of trained immunity in preventing COVID-19 infections and mortality in vaccinated individuals and on community transmission.

## Introduction

Trained immunity, or long-term boosting of innate immune responses, by live vaccines [Bacillus Calmette–Guérin (BCG), measles-containing-vaccine (MCV)] can induce heterologous protection against other pathogens including RNA viruses [1, 2]. The concept of trained immunity has been established by animal/human experimental studies as well as epidemiological studies. It has been speculated that widely administered vaccines could be an important tool for reducing susceptibility to and severity of severe acute respiratory syndrome coronavirus 2 (SARS-CoV-2) infection [2]. One hypothesis is that heterogeneity of mortality rates among different age groups and different countries may be explained to some extent by differing degrees of trained immunity from vaccines that could provide some basal level of protection from this novel pathogen.

Several ecological studies have evaluated the impact of BCG on Coronavirus disease 2019 (COVID-19) outcomes (case incidence, death and case fatality ratio) [3-5]. Results are conflicting, likely due to insufficient control of important factors, including differential timing of the epidemic by country, vaccine coverage, and other health metrics. If live pathogen vaccinations are associated with decreased risk of poor COVID-19 outcomes on a country basis, one might postulate that vaccine coverage could be beneficial both through direct effects on individuals as well as indirectly by decreasing community viral load and therefore transmission to others [2, 6]. We used publicly available national-level data from 140 countries to investigate whether countries with higher proportion of people who received childhood live vaccines (BCG or MCV) have reduced COVID-19 mortality or fewer cases, adding new considerations to previously published analyses.

## Methods

### Data source

We acquired data on COVID-19 through 13 July 2020 from the publicly available databases, Our World In Data (ourworldindata.org) for COVID-19 statistics and Foundation for Innovative New Diagnostics (https://www.finddx.org/at-a-glance/) for COVID-19 testing rates, and standardized them by population and stage of the epidemic [7, 8]. Data on BCG and MCV coverage from 1980 to 2018 were extracted from WHO/UNICEF website [9].

### Outcomes and exposures of interest

We defined the start of the epidemic in each country as the date that country reported a total of 100 cases, and evaluated cumulative deaths and cases reported during the first 60 days of each epidemic [4]. Proportion of vaccinated people in the entire country each year was calculated by multiplying proportion of vaccinated people in the eligible population by estimated proportion of vaccine eligible people (population under age 5) [8]. With vaccine coverage from each year, we calculated a cumulative vaccine coverage index in each country from year XXXX to 2018 (BCG and MCV coverage indices), where we considered XXXX to be 2005 (during the last 15 years), 1995 (during the last 25 years), 1985 (during the last 35 years) and 1980 (during the last 40 years). BCG vaccine coverage data were supplemented by policy of universal vaccination each country with which we assigned zero BCG vaccine coverage to countries each year for which universal vaccination was not implemented (http://www.bcgatlas.org/) [10-12]. We created vaccine indices with different cutoff years to account for duration of trained immunity effect. As expected, number of countries with complete vaccine data with older cutoff (e.g. vaccine indices 1980) is smaller than that with more recent cutoff (2005). We chose BCG/MCV indices 2005 (during the last 15 years) as our exposure vaccine variables based on their predictive abilities for the primary outcome (log-transformed COVID-19 related deaths per million) in bivariate models. However, the BCG and MCV indices from 2005 were found to be highly correlated with these indices from a prior interval (1980) (**Figure 1**).

**Figure 1.**
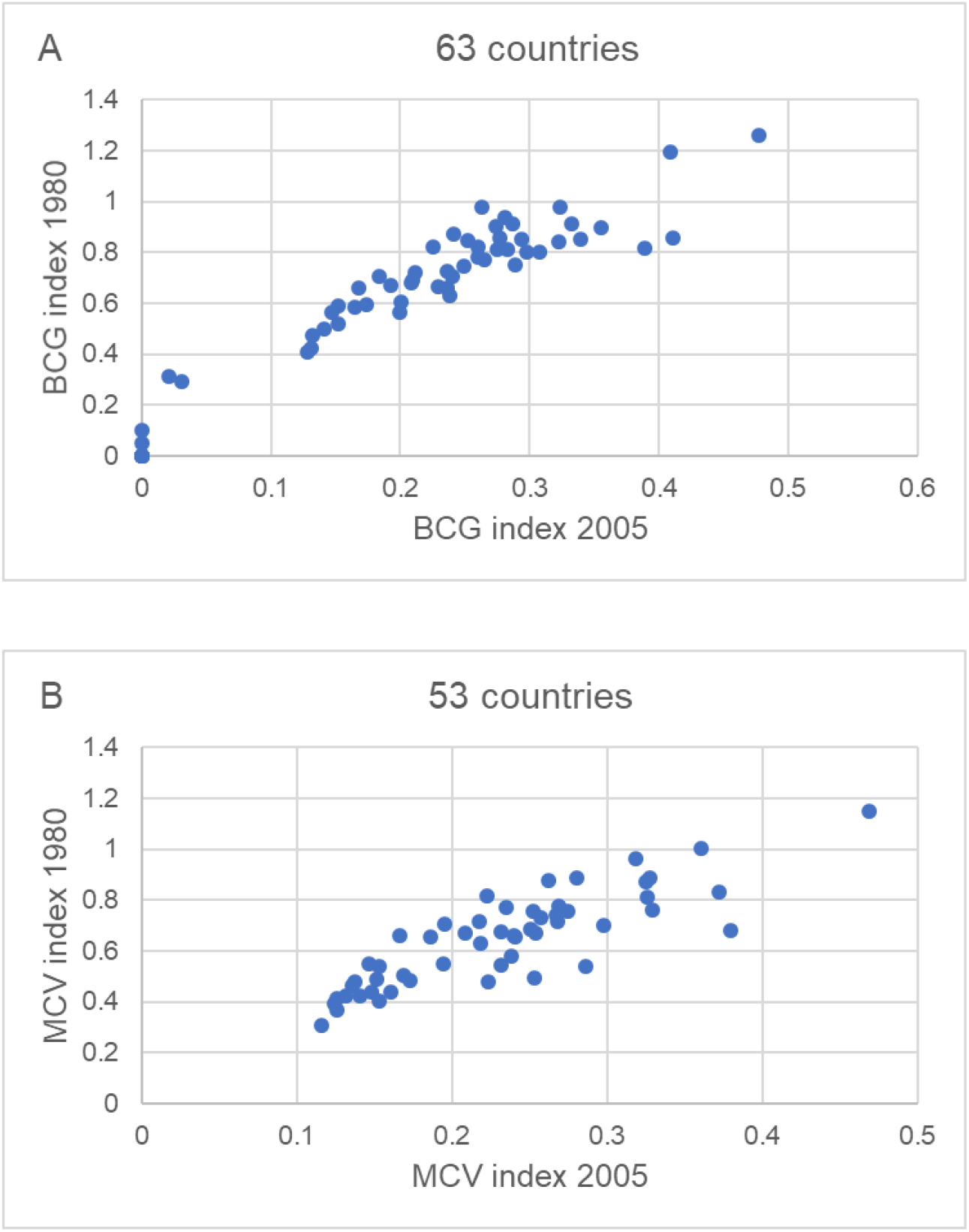
Relationships between vaccine indices 2005 and indices 1980. Scatterplots depict the relationship of each vaccine index by year (2005 vs. 1980) among countries relevant data available. (A) BCG index 2005 vs. BCG index 1980 (R2 = 0.96), (B) MCV index 2005 vs. MCV index 1980 (R2 = 0.85).

### Analysis

Our analysis sought to infer (i) if COVID-19 related death was associated with vaccination exposure variables after controlling for known national characteristics of healthcare systems including life expectancy, prevalence of diabetes, number of hospital beds or physicians per population, COVID-19 testing rate, gross domestic product, composite measure of public health policies, and (ii) whether the association between vaccination exposure and COVID-19 deaths differed as a function of these known national-level variables. Due to small sample size (n=140 countries), we used the Healthcare Access and Quality Index (HAQI), as a single additional predictor in our models since this validated summary score of healthcare system is well correlated with these variables [13, 14].

We included all countries with data on vaccine coverage, COVID-19 outcomes, and the variables comprising the HAQI available to conduct multiple linear regression models. We standardized our vaccine exposure variables as well as the HAQI to have a mean of zero and standard deviation of

1. We used nested linear models to explore:
  i. whether each of the BCG or MCV index is associated with COVID-19 related deaths conditional on HAQI.
  ii. whether BCG and MCV indices are simultaneously associated with COVID-19 related deaths conditional on HAQI.
  iii. whether HAQI modifies the association between both BCG and MCV indices with COVID-19 related deaths.
  iv. whether HAQI modifies the association between BCG index and COVID-19 related deaths.
  v. whether MCV index modifies the association between BCG index and COVID-19 related deaths.

## Results

One hundred forty countries had publicly available complete data with a wide range in COVID-19 cumulative cases (3.3 to 9862.2 per million people) and mortality (0 to 721.2 per million people) 60 days after epidemic start (**Table 1**). Variation of BCG coverage (BCG index 2005 with 136 countries: median 0.24, range 0 to 0.5) is wider than that of MCV coverage (MCV index 2005 with 138 countries: median 0.23, range 0.11 to 0.47). Wide variation of HAQI among 140 countries was also observed (median 68.1, range 18.6 to 97.1).

**Table 1.** Country characteristics and COVID-19 related outcomes at day 60 following epidemic start in each country (N=140)

| **Variable** | **Number of Countries** | **Mean (SD)** | **Median [min, max]** |
| --- | --- | --- | --- |
| Healthcare access and quality index in 2016 | 140 | 63.2 (22.75) | 68.1 [18.6, 97.1] |
| BCG index 2005 (BCG2005) <sup>a</sup> | 136 | 0.23 (0.14) | 0.24 [0, 0.5] |
| MCV index 2005 (MCV2005) <sup>b</sup> | 138 | 0.23 (0.09) | 0.23 [0.11, 0.47] |
| Cumulative COVID-19 deaths/million population <sup>c</sup> | 140 | 48.43 (115.94) | 8.47 [0, 721.16] |
| Cumulative COVID-19 cases/million population <sup>c</sup> | 140 | 1075.17 (1518.79) | 400.24 [3.33, 9862.16] |
| Cumulative COVID-19 deaths (log per million) <sup>c</sup> | 140 | 2.4 (1.68) | 2.19 [-0.69, 6.58] |
| Cumulative COVID-19 cases (log per million) <sup>c</sup> | 140 | 5.98 (1.62) | 5.99 [1.2, 9.2] |

Abbreviations: BCG, Bacillus Calmette–Guérin; MCV, Measles-containing-vaccine; SD, standard deviation.
<sup>a</sup> Estimated proportion of vaccinated people with BCG in the country during the last 15 years.
<sup>b</sup> Estimated proportion of vaccinated people with MCV in the country during the last 15 years.
<sup>c</sup> Cumulative death or case rate for the 60 days since beginning of epidemic in each country, classified as day when 100 total cases had been recorded. Windows at risk differ for each country depending on epidemic start, and were censored at 7/13/20.

BCG was marginally associated with reduced reported COVID-19 deaths with the association remaining after adjusting for HAQI (**Table 2**). However, the magnitude of the association between BCG and COVID-19 deaths increased as a function of HAQI as reflected by a statistically significant interaction term between the BCG vaccine variable and HAQI. Similar patterns were observed for BCG in multivariable models that included both vaccine exposure variables. MCV was also significantly associated with reduced reported COVID-19 death rates, but this association was no longer significant after adjusting for HAQI. MCV did not notably change the association between BCG and COVID-19 deaths. Neither BCG nor MCV coverage were associated with reported COVID-19 cases after adjusting for HAQI (not shown).

**Table 2.**
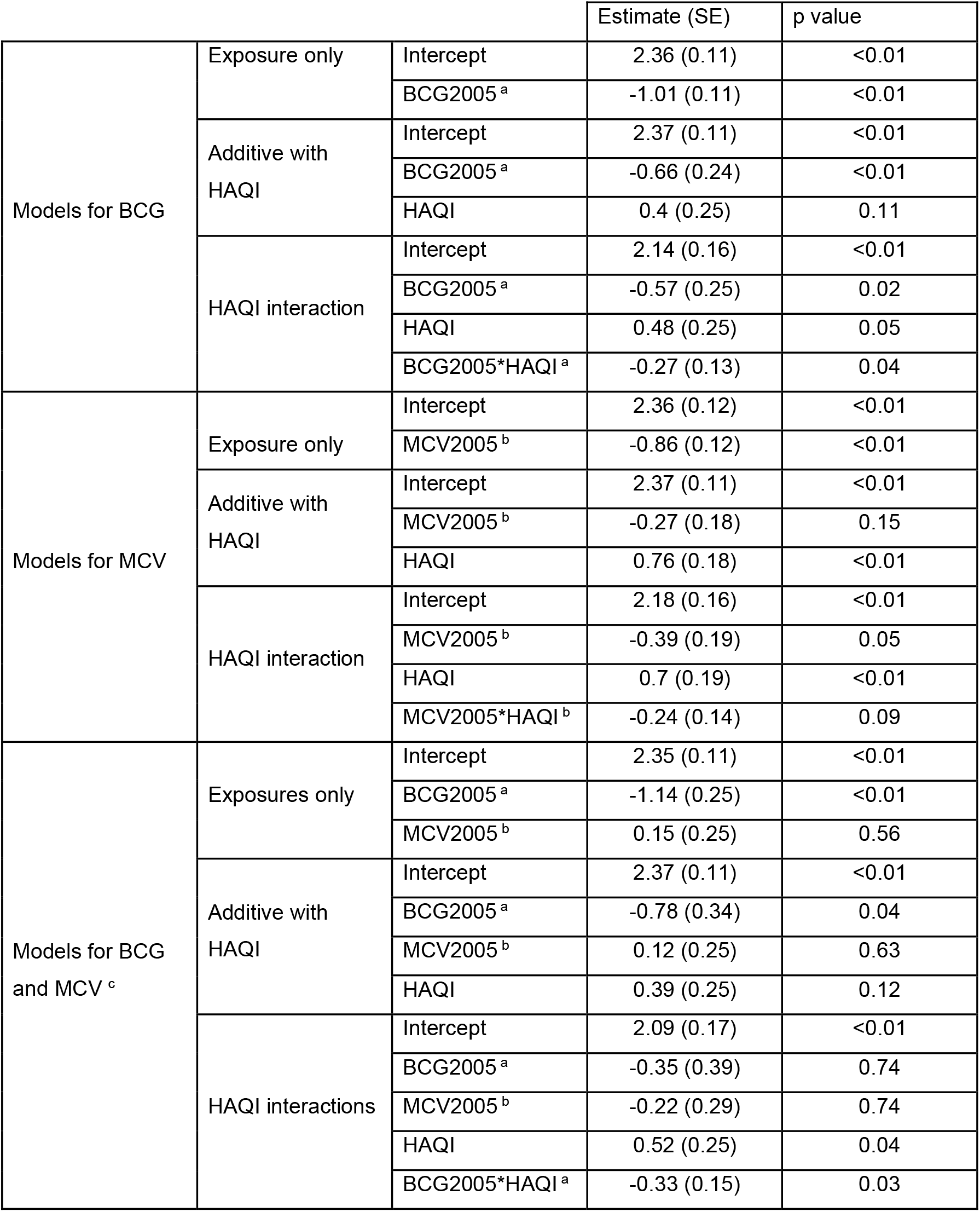
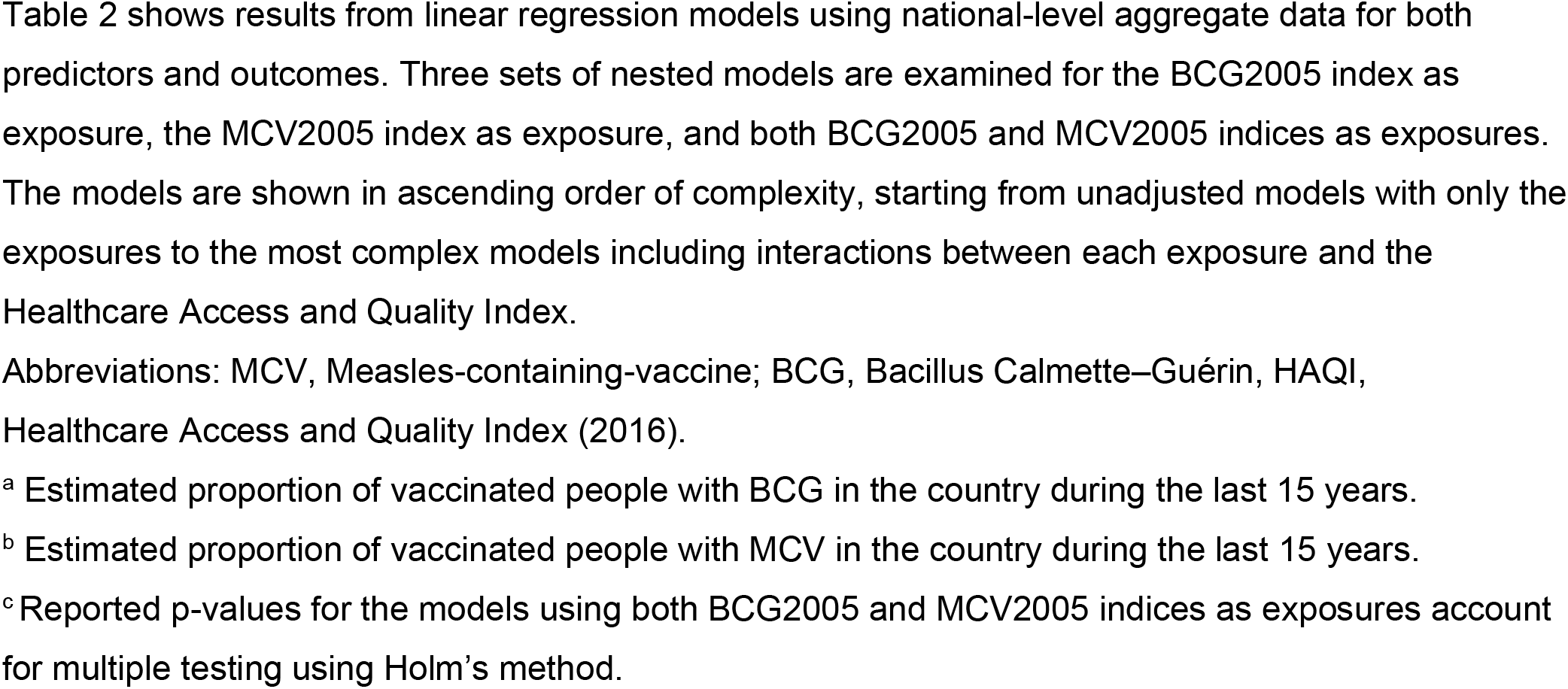
Associations between vaccine coverage and COVID-19 mortality (total COVID-19 related deaths per million at day 60 following epidemic start)

|  |  |  | Estimate (SE) | p value |
| --- | --- | --- | --- | --- |
| Models for BCG | Exposure only | Intercept | 2.36 (0.11) | <0.01 |
|  |  | BCG2005<sup>a</sup> | -1.01 (0.11) | <0.01 |
|  | Additive with HAQI | Intercept | 2.37 (0.11) | <0.01 |
|  |  | BCG2005<sup>a</sup> | -0.66 (0.24) | <0.01 |
|  |  | HAQI | 0.4 (0.25) | 0.11 |
|  | HAQI interaction | Intercept | 2.14 (0.16) | <0.01 |
|  |  | BCG2005<sup>a</sup> | -0.57 (0.25) | 0.02 |
|  |  | HAQI | 0.48 (0.25) | 0.05 |
|  |  | BCG2005*HAQI<sup>a</sup> | -0.27 (0.13) | 0.04 |
| Models for MCV | Exposure only | Intercept | 2.36 (0.12) | <0.01 |
|  |  | MCV2005<sup>b</sup> | -0.86 (0.12) | <0.01 |
|  | Additive with HAQI | Intercept | 2.37 (0.11) | <0.01 |
|  |  | MCV2005<sup>b</sup> | -0.27 (0.18) | 0.15 |
|  |  | HAQI | 0.76 (0.18) | <0.01 |
|  | HAQI interaction | Intercept | 2.18 (0.16) | <0.01 |
|  |  | MCV2005<sup>b</sup> | -0.39 (0.19) | 0.05 |
|  |  | HAQI | 0.7 (0.19) | <0.01 |
|  |  | MCV2005*HAQI<sup>b</sup> | -0.24 (0.14) | 0.09 |
| Models for BCG and MCV<sup>c</sup> | Exposures only | Intercept | 2.35 (0.11) | <0.01 |
|  |  | BCG2005<sup>a</sup> | -1.14 (0.25) | <0.01 |
|  |  | MCV2005<sup>b</sup> | 0.15 (0.25) | 0.56 |
|  | Additive with HAQI | Intercept | 2.37 (0.11) | <0.01 |
|  |  | BCG2005<sup>a</sup> | -0.78 (0.34) | 0.04 |
|  |  | MCV2005<sup>b</sup> | 0.12 (0.25) | 0.63 |
|  |  | HAQI | 0.39 (0.25) | 0.12 |
|  | HAQI interactions | Intercept | 2.09 (0.17) | <0.01 |
|  |  | BCG2005<sup>a</sup> | -0.35 (0.39) | 0.74 |
|  |  | MCV2005<sup>b</sup> | -0.22 (0.29) | 0.74 |
|  |  | HAQI | 0.52 (0.25) | 0.04 |
|  |  | BCG2005*HAQI<sup>a</sup> | -0.33 (0.15) | 0.03 |

Table 2 shows results from linear regression models using national-level aggregate data for both predictors and outcomes. Three sets of nested models are examined for the BCG2005 index as exposure, the MCV2005 index as exposure, and both BCG2005 and MCV2005 indices as exposures. The models are shown in ascending order of complexity, starting from unadjusted models with only the exposures to the most complex models including interactions between each exposure and the Healthcare Access and Quality Index.
Abbreviations: MCV, Measles-containing-vaccine; BCG, Bacillus Calmette–Guérin, HAQI, Healthcare Access and Quality Index (2016).
<sup>a</sup> Estimated proportion of vaccinated people with BCG in the country during the last 15 years.
<sup>b</sup> Estimated proportion of vaccinated people with MCV in the country during the last 15 years.
<sup>c</sup> Reported p-values for the models using both BCG2005 and MCV2005 indices as exposures account for multiple testing using Holm’s method.

We performed additional univariate analyses with COVID-19 testing rate and our other outcomes and exposures to elucidate relationships between COVID-19 metrics and other country-level health and development metrics (**Figure 2**). COVID-19 testing rate was positively correlated with HAQI (R2 = 0.69). COVID-19 testing rate and other markers for better health infrastructure (life expectancy, number of hospitals per population) were negatively correlated with BCG index 2005 (R2 = −0.66, −0.79, −0,51, respectively) and MCV index 2005 (R2 = −0.62, −0.67, −0.58, respectively). Furthermore, COVID-19 testing rate and life expectancy were positively correlated with COVID-19 related mortality (R2= 0.47 and 0.53, respectively). These relationships of healthcare metric components with vaccine coverage and COVID-19 related mortality, suggest that several healthcare system metrics (frequency of COVID-19 testing, life expectancy) are inversely associated with vaccine coverage but positively associated with reported Covid-19 deaths.

**Figure 2.**
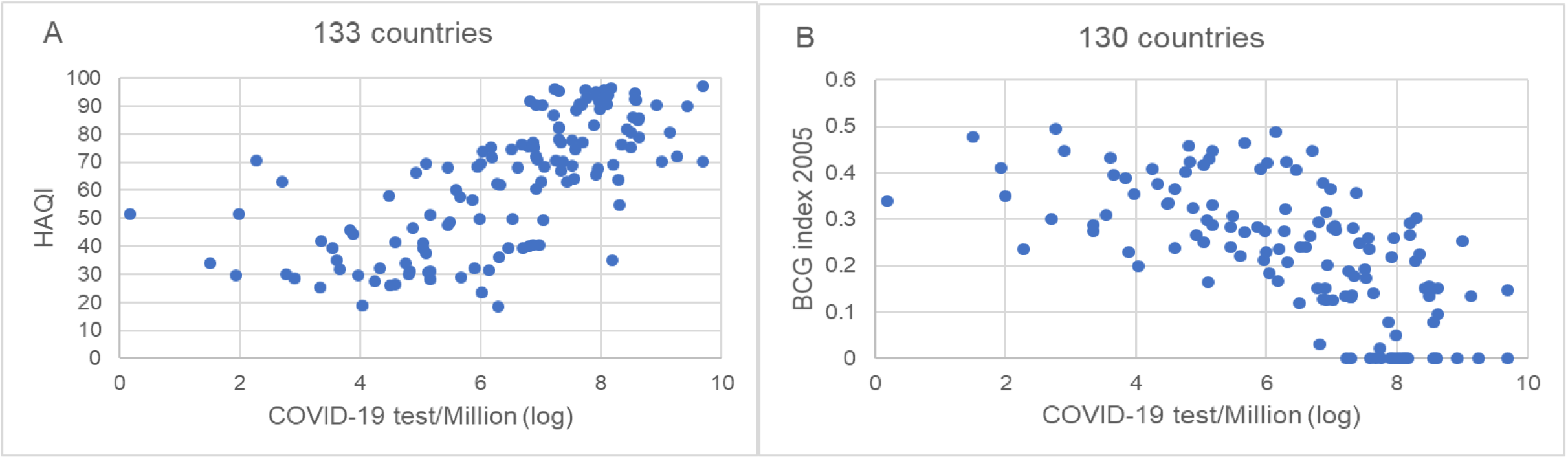
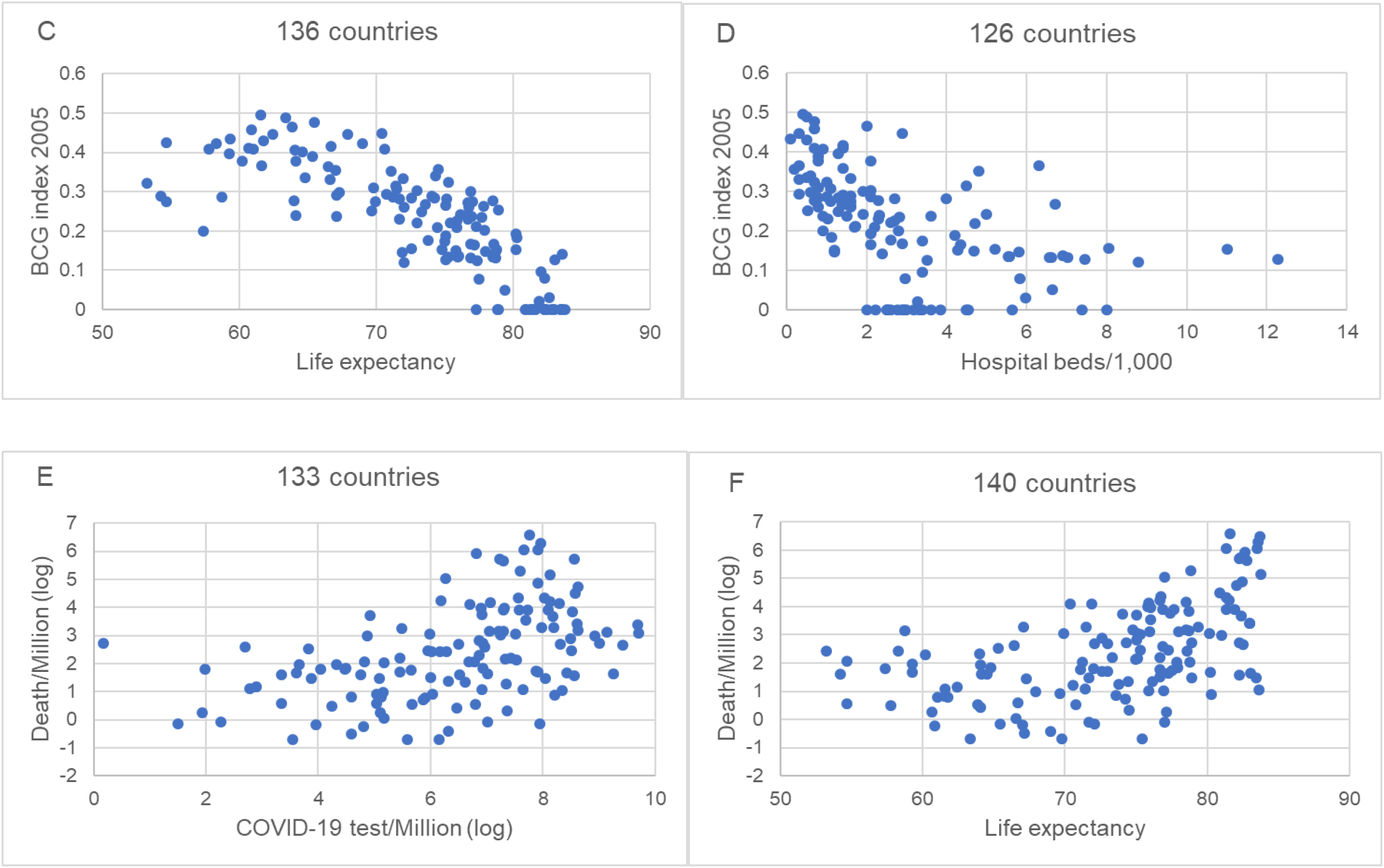
Relationship of healthcare metric compoents with Healthcare access and quality index (HAQI), BCG index 2005 and COVID-19 related mortality. Scatterplots depict the relationship between healthcare metric components and following variables among countries relevant data available. (A) Cumulated total number of COVID-19 testing per million (log-formation) at day 60 following epidemic start each country vs. HAQI (R2 = 0.69), (B) Cumulated total number of COVID-19 testing per million (log-formation) at day 60 following epidemic start each country vs. BCG index 2005 (R2 = −0.66), (C) Life expectancy at birth in 2019 vs. BCG index 2005 (R2 = −0.79), (D) Hospital beds per 1,000 people, most recent year available since 2010 vs. BCG index 2005 (R2 = −0.51), (E) Cumulated total number of COVID-19 testing per million (log-formation) at day 60 following epidemic start each country vs. cumulated total number of COVID-19 related deaths per million (log-formation) at day 60 (R2 = 0.47), (F) Life expectancy at birth in 2019 vs. cumulated total number of COVID-19 related deaths per million (log-formation) at day 60 following epidemic start each country (R2 = 0.53).

## Discussion

MCV and BCG vaccination have demonstrated benefits on reducing childhood mortality in multiple randomized controlled trials (RCTs), the mechanism of which is thought to be trained immunity [15]. The emergence of SARS-CoV-2 heightened the interest and led to several ecological studies to investigate the association between BCG vaccines and COVID-19 related outcomes with conflicting results. Some studies did not account for timing of epidemics in evaluating outcomes and different healthcare system each country, which might have contributed to inconsistent interpretation [3-5]. Although it remains unclear how long the trained immunity effect could last, the design of many prior studies relied on estimated assumptions for its duration or unclear assumptions (e.g. the vaccine variable was whether universal vaccine policy existed or not) [2, 4, 5, 16, 17].

The strengths of this analysis include use of same relative ascertainment window for COVID-19 outcomes for each country considering differential timing of epidemics. Outcome measures at day 60 may also mitigate the issue of delayed report for some countries. We selected deaths as a primary endpoint as a downstream parameter to account for all possible vaccine effects (preventing acquisition of the virus, symptomatic episodes, severe illness, or transmission by accelerating viral clearance) [1, 2, 17-19]. Our models included BCG, MCV and HAQI considering effect modification and confounding [14]. Although we selected BCG/MCV 2005 indices since these had the strongest association in bivariate analysis, quantifying the cumulative vaccine exposure for each vaccine over the last 15 years, the 2005 and 1980 indices for each vaccine were also highly correlated (**Figure 1**); these indices therefore allow for extrapolation to vaccine coverage for a larger proportion of the middle-age and older population [20].

The current study demonstrated marginal associations between coverage of both vaccines with COVID-19 related mortality. The significance of BCG coverage remained in the conditional model after controlling for HAQI. The association of MCV coverage with COVID-19 deaths was weaker than that of BCG coverage in all models. This may reflect lower variability of MCV coverage between countries or indicate this is a surrogate marker for other factors associated with COVID-19 outcomes if not a biologic difference. Somewhat counter-intuitively, COVID-19 related mortalities were higher in countries with higher HAQI. Since countries with higher HAQI also were more likely to have higher COVID-19 test rates (**Figure 2**), this likely describes a phenomenon of under ascertainment of both cases and deaths in countries with less health infrastructure, or else COVID-19 containment strategies not reliant on testing (masking, social distancing). We chose not to undertake a more complicated multivariable analysis given the small sample size (n=140) and compounding of uncertainty within each country-level metric. Interestingly, the magnitude of the association between BCG coverage and COVID-19 deaths increased as a function of HAQI. This suggests either that there is a true relationship between BCG coverage and COVID-19 outcomes masked by health-system heterogeneity, or else the association could be attributed to an unmeasured confounder correlated with BCG coverage but not HAQI, such as confidence in the government or low parity in access to care despite adequate resources [21]. Because the majority of countries with high HAQI no longer use BCG, sensitivity analyses focusing on high-HAQI countries were not suitable.

We explicitly acknowledge the principal limitation that country-level data does not represent individual exposures or outcomes [22]. There are other potential confounders such as genetic variance, national COVID-19-specific public health responses (e.g. percent mask use, policy and adherence to movement restrictions), heterogeneity in population density and disparities in access to care, the majority of which are difficult to quantify [5, 21, 23]. Although studies have indicated certain BCG strains might induce more effective trained immunity than others, there was insufficient data to evaluate these hypotheses [3, 24]. Moreover, our analysis treated the national-level summaries as fixed variables, ignoring the uncertainty in these estimates and possibly their biases. The p-values reported in **Table 2** are thus under-estimates of the true p-values. Finally, our models only examined linear associations.

In conclusion, we found an association between higher cumulative BCG coverage and COVID-19 related deaths but not cases, and only a marginal effect of MCV coverage on either; we cannot rule out that these observations are attributable to differential healthcare infrastructure, including COVID-19 testing rate and population age distribution or other unmeasured confounders. Several RCTs (primarily BCG) are currently underway (https://clinicaltrials.gov/) to evaluate the impact of live pathogen vaccines on COVID-19 related outcomes. Some of them may however not be able to adequately assess the vaccine efficacy depending on number and types of subjects, primary endpoints, and state of local epidemics [2, 17, 20, 24]. Cluster randomized trials of booster BCG or MCV vaccines could best evaluate both personal and community-level effects of trained immunity.

## Data Availability

Original data are available by request to the corresponding author.

## Acknowledgments

We thank Janet A. Englund for funding support and Elizabeth M. Krantz for analytic input.

